# Histological characteristics of the quadriceps muscle and their associations with gait in older adults

**DOI:** 10.64898/2026.11.10.26367782

**Authors:** Shahram Oveisgharan, Savannah Longo, Jingyun Yang, Julie A. Schneider, David A. Bennett, Osvaldo Delbono, Aron S. Buchman

**Affiliations:** Rush Alzheimer’s Disease Center, Rush University Medical Center, Chicago, IL, USA; Department of Neurological Sciences, Rush University Medical Center, Chicago, IL, USA; Department of Internal Medicine, Gerontology and Geriatric Medicine. Wake Forest University School of Medicine, Winston-Salem, NC, USA; Department of Pathology, Rush University Medical Center, Chicago, IL, USA

## Abstract

**Background/objective:** There is a paucity of data about muscle morphology in aging adults. We quantified histologic features of the quadriceps muscle and examined their associations with gait function and physical activity in community-dwelling older adults.

**Methods:** Participants were from the Rush Memory and Aging Project. Postmortem sections of the quadriceps muscles were examined by immunohistochemistry using antibodies against myosin heavy chain, and histological characteristics of the quadriceps muscles were derived through images quantifications and segmentations. In addition, brain sections were assessed for collection of Alzheimer’s disease and related dementias pathologies. Before death, participants’ physical activity was objectively measured through wearing an accelerometer sensor for 10 days, and gait function was quantified by walking 8 feet and turning 360° twice. Linear regressions and compositional analyses were used for analyzing data.

**Results:** Postmortem quadriceps was examined in 236 decedents (mean age at death was 91.7±6.0 years, and 71.6% (n=169) were females. On average, 44.7% (SD=19.5) of the muscle fibers were type I and 54.2% (SD=19.4) were type II, and 40.6% (SD=13.0) and 21.9% (12.9) of the quadriceps muscle area were composed of type I and II muscle fibers, respectively. In compositional analyses, male sex (estimate=0.303, SE=0.126, p=0.016) and more physical activity (estimate=0.118, SE=0.059, p=0.045) were associated with larger quadriceps muscle areas with type II muscle fibers, which were related to better gait (estimate=-0.049, SE=0.021, p=0.021). Quadriceps muscle composition explained an additional 3.2% of gait’s variance beyond the 5.8% variance explained by Alzheimer’s disease and related dementias brain pathologies.

**Conclusion:** Quadriceps muscles having larger areas with type II muscle fibers were associated with more physical activity and better gait function in aging adults. Elucidating the molecular mechanisms underlying these associations are critical for developing new therapies.

## INTRODUCTION

Declining gait is common in older adults and leads to loss of independence and diverse adverse health outcomes. As many as one in four older Americans suffer from mobility disability, more common than other disabilities including cognitive disability^1^. Though daily physical activity is thought to be a modifiable risk factor to prevent mobility disability, inexorable mobility decline occurs in most aging adults as the biology and pathologic basis of impaired mobility remains poorly characterized.

While neural control of gait may originate in the brain, motor pathways extend throughout the entire brainstem and spinal cord to exit the central nervous system (CNS) via peripheral nerve to reach the muscle, the final effector of movements. Many prior studies investigating the pathologic basis for motor impairment in older adults have focused on the brain and have shown that Alzheimer’s disease and related dementias (ADRD) pathologies as well as structural degradation of brain white matter are associated with age-related motor decline^2–5^. Yet, there has been longstanding recognition that many older adults show prominent loss of both muscle bulk and function, “sarcopenia”^6,7^, that is related to impaired mobility and adverse health outcomes^8,9^. Although muscle biopsy can be obtained in living older adults, it is invasive, costly, and not routinely available. So, most muscle studies have employed non-invasive imaging techniques, such as magnetic resonance and CT imaging,^10,11^ bioimpedance^12^, or ultrasound^13^, that do not differentiate different histologic muscle components.

Despite the critical role of muscle as a proxy of late life mobility impairment, there are few studies that quantified age-related histological features of muscle in large numbers of well-characterized community-dwelling older adults and examined their associations with muscle function.^14^ ^15^ Moreover, we are unaware of studies that examined the associations of both muscle histology and ADRD brain pathologies with mobility in the same individuals. To fill these knowledge gaps, the current study builds on our prior clinical and postmortem studies^16–19^ of community-dwelling older adults to quantify histological indices of postmortem quadriceps muscle in decedents from whom indices of ADRD brain pathologies had previously been collected. These data were integrated with quantitative metrics of physical activity and gait function that had been collected prior to death.

## METHODS

### Participants

The participants were from the Rush Memory and Aging Project (MAP), an ongoing longitudinal clinical pathologic cohort study recruiting community-dwelling older adults across northeastern Illinois since September 1997. MAP participants live in the community and are enrolled, interviewed, and tested in their residences, including individual homes, retirement centers, and subsidized senior housing. Individuals are enrolled if they do not have known dementia and are willing to consent to annual clinical assessments and autopsy at the time of death for donation of brain and other tissues including the muscle. Staff collecting and analyzing clinical, device-derived, or postmortem data are blinded to other data sources. The current study included the first 236 participants with completed histologic assessment of postmortem quadriceps muscle.

### Histological assessment of the quadriceps muscle

The median interval between death and the autopsy procedures was 7.1 hours (IQR: 6.0–9.0). At the time of autopsy, the rectus femoris of the quadriceps muscles was dissected, and several sections were embedded in optimal cutting temperature (OCT) compound, and frozen in nitrogen-cooled isopentane containers. Samples were stored at -80°C until cut into 10-µm sections at -20°C using a Leica CM3050 cryostat (Leica Biosystems Inc., Buffalo Grove, IL, USA).

To process samples for immunofluorescence, sections were rinsed in Phosphate-Buffered Saline (PBS) and blocked with 10% goat serum in PBS for 1 h. The primary antibodies used for myosin heavy chain (MHC) isoforms were purchased through the University of Iowa Developmental Studies Hybridoma Bank: BA-F8 (MHC-I, 1:50), SC-71 (MHC-IIa, 1:500), and 6H-1 (MHC-IIx, 1:50). To mark the extracellular space, we used MAB1914P anti-laminin gamma 1, clone A5 rat primary antibody (laminin, 1:100) (MilliporeSigma, Burlington, MA, USA). Primary antibodies were incubated at room temperature for 2 h with 10% goat serum in PBS. Sections were washed twice in PBS for 5 min and incubated with secondary goat anti-mouse antibodies for MHC: AlexaFluor 350 IgG2b (1° BA-F8, 1:500), AlexaFluor 488 IgG (1° SC-71, 1:500), and AlexaFluor 555 IgM (1° 6H-1, 1:500). We used goat anti-rat IgG Alexa Fluor 680 (1:500) secondary antibody for laminin. All secondary antibodies were obtained from Invitrogen (Thermo Fisher Scientific Corp., Carlsbad, CA, USA). Slides were mounted using the Dako fluorescence mounting medium (Dako North America, Carpinteria, CA, USA). Slides were stored at 4°C in a microscope slide box until imaged.

#### Image quantification

Muscle cross-sections were imaged using a Hamamatsu C10600-10B-H ORCA-R2 CCD camera (Hamamatsu Corp., Bridgewater, NJ, USA) mounted on an Olympus IX81 inverted fluorescence microscope (Olympus America, Center Valley, PA, USA) at 10× and 20 x magnifications. Fluorescence filter sets (Chroma Technology Corp., Bellows Falls, VT, USA) included 41017 Endow GFP/EGFP Bandpass, 41004 Texas Red, 31000v2 DAPI/Hoechst/AMCA, and 41022 Cy5.5 (Red-Shifted). Individual single-channel images were merged into multichannel composites using MetaMorph Basic software (Molecular Devices, San José, CA, USA).

Image contrast was standardized using a calibrated acquisition and normalization workflow comprising: (i) fixed exposure times for each fluorescence channel, (ii) session-specific flat-field correction, and (iii) uniform histogram-based normalization applied identically to all raw images prior to segmentation. Quality-control thresholds and rejection criteria were defined a priori. To monitor inter-session drift, a designated sentinel sample from the previous imaging batch was re-imaged at each new session. Acceptance criteria required <5% variation in fiber cross-sectional area and no deviation in fiber-type proportions; sessions exceeding these thresholds were recalibrated and re-imaged. This workflow ensured consistent intensity scaling and minimized operator-dependent variability across imaging sessions. Histological analyzers were blinded to both pathology and clinical data.

#### Semiautomatic muscle analysis script

A semiautomated muscle-analysis script was developed within the NIH ImageJ/Fiji environment^20^. After selecting a region of interest (ROI), used to exclude areas unsuitable for analysis such as folds, tears, or inconsistent labeling, the script proceeds through four sequential, user-controlled steps. **(1) Laminin segmentation.** Following ROI selection, the laminin/dystrophin channel is processed automatically. Background subtraction is performed by subtracting a high-radius Gaussian-filtered duplicate from the raw image, followed by global thresholding using the *Triangle* method in ImageJ. The resulting binary image is duplicated, dilated to close small boundary gaps, and skeletonized. The skeleton is then overlaid onto the thresholded image to generate continuous fiber outlines, which are segmented using the Morphological Segmentation tool from the MorphoLibJ plug-in^21^. **(2) Segmentation editing.** Segmented fibers are overlaid onto an RGB composite image for user review. Fibers are added to an ROI list for downstream analysis, and users can edit the segmentation by removing inaccurately segmented fibers or adding fibers missed during automated detection. **(3) Fiber type detection and analysis.** Fiber typing is performed identically for each subtype. After segmentation editing, the relevant fluorescence channel is selected, and background subtraction using a high-radius Gaussian filter is applied. A noise-reduction filter minimizes the influence of bright, nonspecific pixels caused by localized antibody accumulation. The image is then thresholded using the *Triangle* method to identify fiber-type–specific labeling. **(4) Fiber type editing.** Automatically assigned fiber types are displayed as a color-coded map. Users can manually correct fiber type assignments by modifying the color designation of individual fibers directly within the map. The script proceeds sequentially through these steps, allowing user oversight at each stage. Reproducibility, validation, and quality-control features of the script and associated code have been previously reported^20^. **Figure 1** shows a representative immunostained muscle cross-section and the corresponding segmentation, and **Figure e-1** illustrates three muscle samples enriched for different muscle fibers.

**Figure 1.**
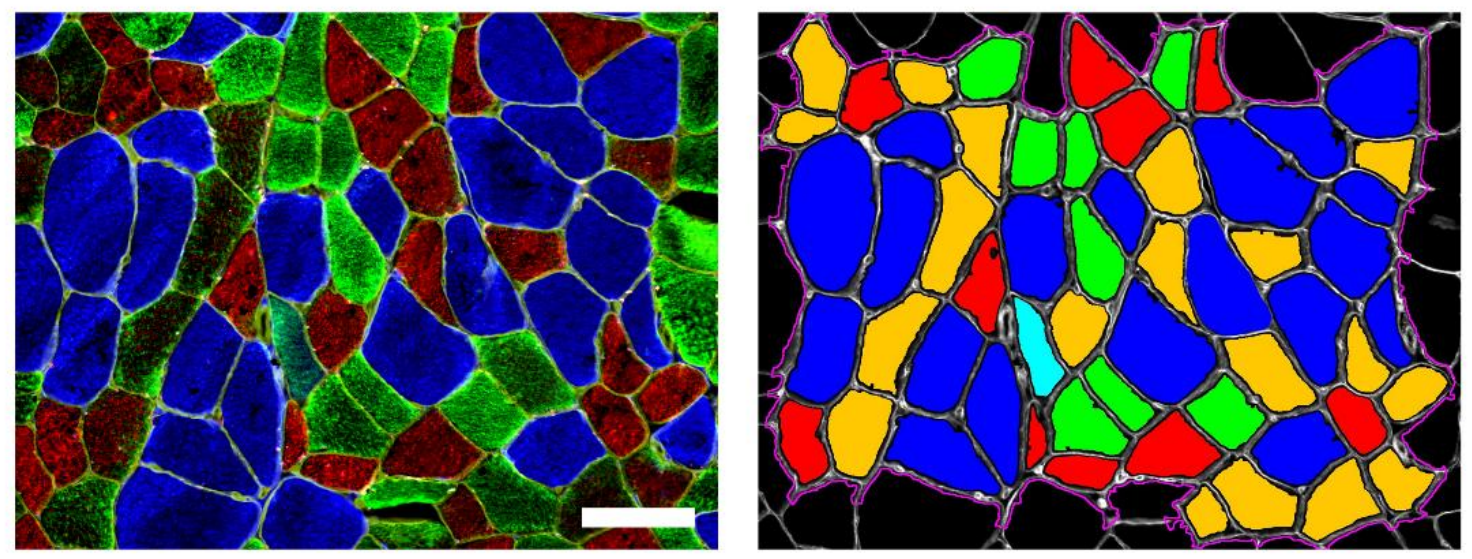
Muscle fibers segmentation. A representative muscle cross-section immunostained for type I (blue), IIA (green), and IIX (red) myosin heavy chains (left) and the corresponding segmentation (right). Type IIA + type I hybrid fibers and type IIA + type IIX hybrid fibers are shown in cyan and orange, respectively. Scale bar = 100 µm. Incomplete fibers at the image borders were excluded from the analysis.

### Postmortem assessment of ADRD brain pathologies

After brain removal, one hemisphere was frozen for future studies, and the other hemisphere was fixed in a 4% paraformaldehyde solution. ADRD pathological indices were obtained by examining the 1-cm slabs and the subsequent sections that were prepared from predetermined regions.

#### Alzheimer’s disease (AD)

Immunohistochemistry and digital image analyses were used to quantify amyloid-β (Aβ) load^22^. The following antibodies were used for staining Aβ: 4G8 dilution 1:9000 (Covance Labs, Madison, WI), 6F/3D dilution 1:50 (Dako North America Inc., Carpinteria, CA), 10D5 dilution 1:600 (Elan Pharmaceuticals, San Francisco, CA). Thin sections were prepared from 8 brain regions: mid frontal, superior frontal, cingulate, inferior temporal, entorhinal, angular gyrus, calcarine cortices, and the hippocampus. The percentages of pixels stained with Aβ were calculated to yield Aβ load. Sections from the same 8 brain regions were used for immunohistochemical quantification and digital image analysis of paired helical filament tau tangles using antibodies against phosphorylated tau (AT8 dilution 1:1000, Innogenetics, Alpharetta, GA). Ratios of tau tangle counts to the examined areas were used to calculate tau tangle density. Quantified neuritic plaques, neurofibrillary tangles (determined by staining with Modified Bielschowsky silver stain), and Thal stages (based on the immunohistochemical staining against Aβ) were used to adjudicate AD pathological diagnosis^22,23^.

#### Limbic-predominant age-related TDP-43 encephalopathy neuropathologic changes (LATE-NC) ^24^

LATE-NC was quantified immunohistochemically using antibodies against phosphorylated transactive response DNA binding protein 43 (TDP-43; pS409/410 1:100). The examined brain regions were the amygdala, hippocampus (CA1, subiculum), dentate gyrus, entorhinal cortex, and neocortices (orbital frontal, midfrontal, anterior temporal, and middle temporal cortices). In the current study, LATE-NC was summarized using a dichotomous variable indicating the presence of TDP-43 inclusions in the entorhinal cortex, hippocampus, or beyond.

#### Hippocampal sclerosis^25^

Hippocampal sclerosis was present if severe neuronal loss and gliosis were observed in the CA1 and subiculum regions of the hippocampus.

#### Parkinson’s disease (PD) pathology

Lewy bodies were detected through immunohistochemistry using antibodies against phosphorylated alpha-synuclein protein (Zymed LB 509 dilution 1:50; pSyn dilution 1:20 000; Wako Chemicals)^26^. The examined brain regions were midfrontal, midtemporal, inferior parietal, anterior cingulate, entorhinal, hippocampus, basal ganglia, and midbrain. Nigral neuronal loss was assessed in the midbrain sections at the level of the third cranial nerve exit root containing substantia nigra. PD pathology was present if Lewy bodies were detected and moderate to severe nigral neuronal loss was present^27^.

#### Macroinfarcts

Fixed slabs were examined for infarcts, which were confirmed under microscope. Chronic macroinfarcts, characterized by cavities surrounded by few macrophages and chronic gliosis, were quantified and summarized by a dichotomous variable indicating the presence of one or more chronic macroinfarcts^28^.

#### Microinfarcts

Nine brain regions were searched for chronic microinfarcts, characterized as scar-like lesions without cavities or as pial surface invagination. The 9 brain regions were six cortical brain regions (frontal, temporal, entorhinal, hippocampal, parietal, and anterior cingulate), two subcortical regions (anterior basal ganglia and thalamus), and the midbrain. A dichotomous variable was used to indicate one or more chronic microinfarcts^29^.

#### Atherosclerosis

Vessels of the Circle of Willis and their proximal branches were examined for atherosclerosis and the consequent lumen narrowing, which were summarized with a dichotomous variable indicating moderate to severe atherosclerosis^30^.

#### Arteriolosclerosis

Small arteries and arterioles in the anterior basal ganglia region were examined for arteriolosclerosis, vessel wall thickening and lumen narrowing, which were summarized using a dichotomous variable indicating moderate to severe arteriolosclerosis^31^.

#### Cerebral amyloid angiopathy

Immunohistochemistry using antibodies against Aβ, as described above, was used to visualize Aβ deposits in the meningeal and parenchymal vessel walls in four brain regions (frontal, temporal, calcarine, and angular cortex). The severity of Aβ deposits was summarized using a dichotomous variable indicating moderate to severe cerebral amyloid angiopathy^32^.

### Assessment of physical activity

To circumvent the limitations of self-report history of physical activity, including recall bias, we objectively measured physical activity using an accelerometer sensor (Actical®; Philips Healthcare, Bend, OR, USA) worn over the non-dominant wrist for 10 days. Activity counts of 15-second epochs were summed for each 24-hour period of complete recording and averaged across the wear days to yield total daily physical activity counts as described in previous publications^33^. To contextualize this measure of physical activity, prior studies reported activity counts recorded by sensors during varied physical performances in older adults. For example, walking at 2.5 mph is associated with 2354 activity counts/min^34^.

### Assessment of gait function

At annual visits, the participants were asked to walk 8 feet and turn 360° twice. The reciprocal of time and the number of steps to complete each of these tasks were used to derive a summary measure of gait function supported by previously published principal component analysis^35^. Lower values of this summary gait function measure indicate slower gait and more steps and were associated with ADRD brain pathologies in our prior publications^17,19^.

### Assessment of other clinical covariates

Age at death was calculated from date of birth, obtained at the MAP baseline visit, and date of death. Sex, race, and ethnicity were also obtained at baseline.

Self-report questions were used to assess vascular risk factors (hypertension, diabetes, smoking) and vascular diseases (stroke, myocardial infarction, heart failure, and lower extremities claudication), which were summarized as composite variables indicating the number of conditions present. Participants were also asked lower back pain and pain in the hip, knee, or feet that had lasted at least one month and had been present in most days. Body mass index was calculated as using weight and height (weight(kg)/height(m)^2^) measured in the visits.

Participants completed 21 cognitive tests^28^, and dementia diagnosis was assigned by dementia-expert clinicians, after reviewing the cognitive tests’ score and select clinical data, based on published criteria^36^.

### Statistical analysis

Separate linear regression models were used to examine the associations of age at death and sex (Model1) and physical activity (Model 2) with the muscle histological characteristics. Each characteristic was examined in a separate model. To examine associations of individual histological characteristics with gait, we used separate linear regression models with gait function as the outcome and one histological characteristic as the model term adjusting for age at death, sex, and the time interval between last gait assessment and death.

Yet, muscle is composed of different components including different muscle fibers, connective tissue, and blood vessels. Because higher values of one component are accompanied by lower values of other components, these components are not independent to be examined together in a single linear regression model. So, we employed an analytic approach, compositional analyses, that has been developed to accommodate this interdependence and yield interpretable inferences when the components are analyzed together. Compositional analysis has been used to examine diverse scientific and clinical data^37,38^. To perform compositional analysis, we created ratios, by dividing the area of each muscle component by the sum of all the components, and analyzed these ratios because the sum of the ratios equaled 1 (or 100%), a criterion required for using compositional data analysis^39^.

For parsimony, we summarized the examined quadriceps muscle sections by defining four components: the muscle area composed of type I muscle fibers, type II muscle fibers, mixed type I-II muscle fibers, and non-muscle fiber tissues (including blood vessels, connective tissues, and fat). For example, an average participant had 40% of the examined area composed with type I muscle fibers, 22% with type II muscle fibers, 1% with mixed type I-II muscle fibers, and 37% with non-muscle fiber tissues. Because we did not observe mixed muscle fibers in 34.3% (n=81) of the quadriceps samples, and none of the components can be zero in a compositional analysis, we replaced zeros with the minimum of non-zero mixed muscle fiber area ratio, which was 0.000075. Then, we reduced 0.000075 from the ratios of type I and type II muscle fibers. For example, if type I, II, mixed type I-II, and non-muscle fibers areal ratios were 0.40, 0.22, 0, and 0.38, respectively, we replaced the composition with the following: type I=0.399952, type II=0.219973, mixed type I-II=0.000075, non-muscle fiber=0.38.

First, we did compositional analyses to examine the associations of age at death, sex, and physical activity as the predictors with the four muscle components as the outcome. We chose the muscle area composed of mixed type I-II muscle fibers as the reference component and made three ratios through dividing the areas of type I, type II, and non-muscle fibers by the area of the reference component. Mixed fibers were used as the reference as they were biologically minor, did not privilege type 1 or type II fibers, and allowed interpretation of reallocations among functionally meaningful compartments. The three ratios were log-transformed and were examined as the outcome of two linear mixed-effects models. Predictors for these models included age at death, sex (Model 1) and physical activity (Model 2), group (a 3-level categorical variable corresponding to the three ratios), and interaction of the group with the other terms. A random term for intercept was also included in the model. To illustrate the findings, we used ternary plots that illustrate simplex surfaces where the coordinate axes are constrained to sum 1^39^. Additional mixed effects models further adjusted for vascular risk factors and diseases, body mass index, low back and lower extremities’ joints pain, dementia and interaction of these variables with the group to assess whether the association of physical activity with muscle composition persisted after accounting for these possible confounders. All analyses involving physical activity also adjusted for the time interval between the last physical activity measurement and death.

Second, we used a compositional isotemporal substitution model^40^ to examine reallocations between the four components in relation to gait as the outcome. Three variables were computed from the four components of the muscle, type I, II, mixed I-II, and non-muscle fibers areas, with the mixed type I-II muscle fibers as the reference category. The three computed variables were^40^:

*Variable 1*= ln(square root(area of type I muscle fibers×area of type II muscle fibers)/square root(area of mixed muscle fibers×non-muscle fibers area)).

*Variable 2*= 0.5^1/2^×ln(area of type I muscle fibers/area of type II muscle fibers).

*Variable 3*= 0.5^1/2^×ln(area of mixed muscle fibers/area of non-muscle fibers).

We used a linear regression model to examine the association of these three variables as the predictors with gait function as the model outcome, adjust for age at death, sex, and the time interval between gait assessment and death. Then, we estimated the change in gait associated with the change between two compositions: a reference composition (approximately the means of the four muscle areas: area composed of type I muscle fibers (40%), type II muscle fibers (22%), mixed type I-II muscle fibers (1%), and non-muscle fibers (37%)) and a new composition created by reallocation from one component of the composition to another. For example, a new composition was made by reducing 5% of the percentage area of type II muscle fibers (22% -> 17%) and increasing 5% to the percentage area of type I muscle fibers (40% -> 45%). Of note, we used the word reallocation to be consistent with the statistical papers describing compositional isotemporal substitution models^40^, but we do not mean any causal association between the muscle components and gait.

The analyses were done using SAS version 9.4. Ternary plots were generated in R version 4.2.2. Two-sided p-values less than 0.05 were considered statistically significant.

## RESULTS

### Demographic and clinical characteristics

The demographic and clinical characteristics of the analytic cohort (N=236) are summarized in **Table 1**. The mean age at death was 91.7 years (SD=6.0), 71.6% (n=169) females, and 30.3% (n=71) had dementia before death. Mixed brain pathologies were common at autopsy (median of the number of pathologies=3, IQR: 2–4), and the most common pathology was AD present in 66.5% (n=157) of participants.

**Table 1.**
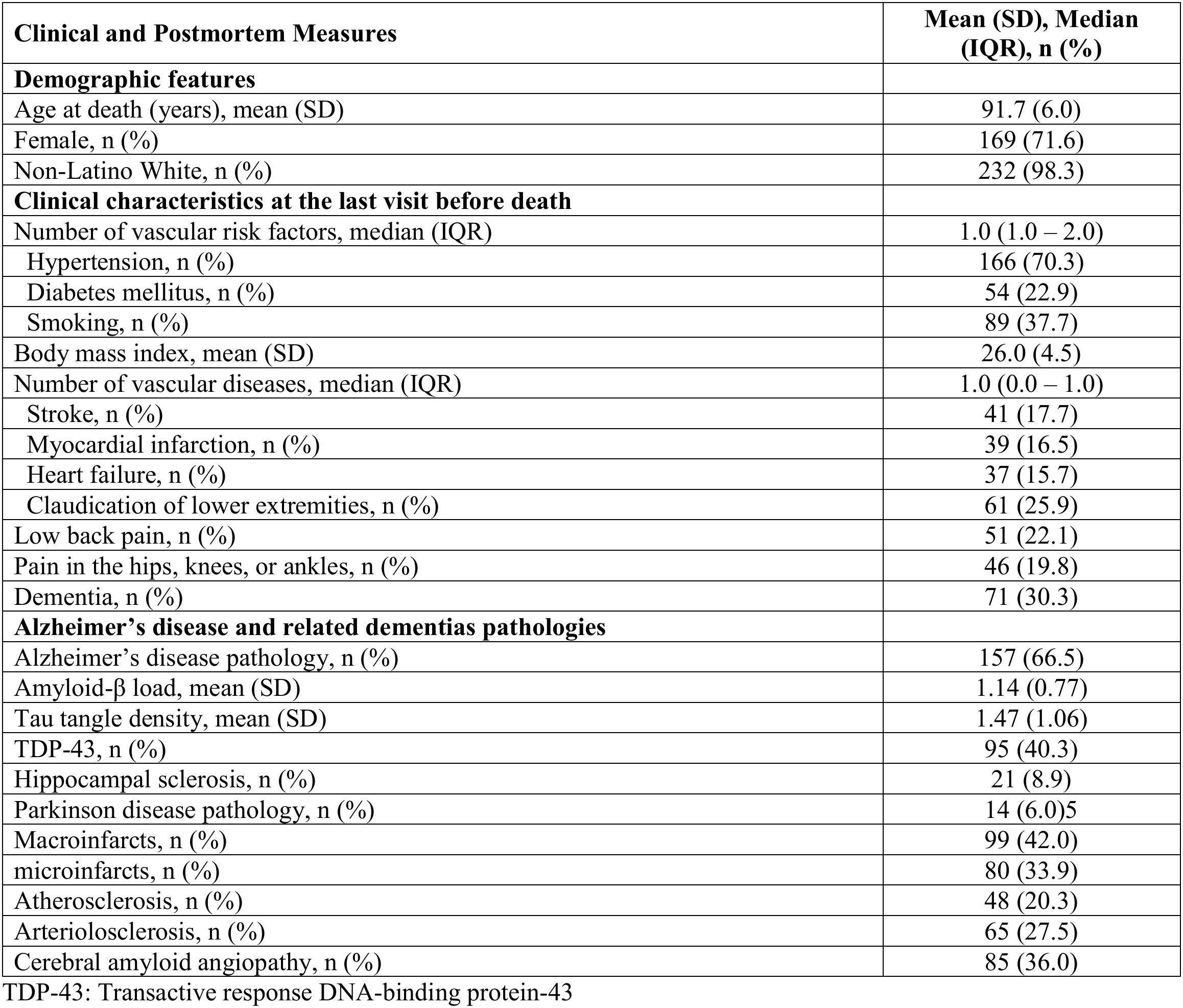
Description of the analytic cohort (N=236).

| **Clinical and Postmortem Measures** | **Mean (SD), Median (IQR), n (%)** |
|---|---|
| **Demographic features** |  |
| Age at death (years), mean (SD) | 91.7 (6.0) |
| Female, n (%) | 169 (71.6) |
| Non-Latino White, n (%) | 232 (98.3) |
| **Clinical characteristics at the last visit before death** |  |
| Number of vascular risk factors, median (IQR) | 1.0 (1.0 – 2.0) |
| Hypertension, n (%) | 166 (70.3) |
| Diabetes mellitus, n (%) | 54 (22.9) |
| Smoking, n (%) | 89 (37.7) |
| Body mass index, mean (SD) | 26.0 (4.5) |
| Number of vascular diseases, median (IQR) | 1.0 (0.0 – 1.0) |
| Stroke, n (%) | 41 (17.7) |
| Myocardial infarction, n (%) | 39 (16.5) |
| Heart failure, n (%) | 37 (15.7) |
| Claudication of lower extremities, n (%) | 61 (25.9) |
| Low back pain, n (%) | 51 (22.1) |
| Pain in the hips, knees, or ankles, n (%) | 46 (19.8) |
| Dementia, n (%) | 71 (30.3) |
| **Alzheimer’s disease and related dementias pathologies** |  |
| Alzheimer’s disease pathology, n (%) | 157 (66.5) |
| Amyloid-β load, mean (SD) | 1.14 (0.77) |
| Tau tangle density, mean (SD) | 1.47 (1.06) |
| TDP-43, n (%) | 95 (40.3) |
| Hippocampal sclerosis, n (%) | 21 (8.9) |
| Parkinson disease pathology, n (%) | 14 (6.0)5 |
| Macroinfarcts, n (%) | 99 (42.0) |
| microinfarcts, n (%) | 80 (33.9) |
| Atherosclerosis, n (%) | 48 (20.3) |
| Arteriolosclerosis, n (%) | 65 (27.5) |
| Cerebral amyloid angiopathy, n (%) | 85 (36.0) |

TDP-43: Transactive response DNA-binding protein-43

### Quadriceps muscle histologic characteristics

Immunohistochemistry using myosin heavy chain staining of the mounted quadriceps sections assessed 3 different muscle fiber types (**Figure 1**) summarized in **Table 2**. Each section was partitioned into areas composed of muscle fibers or non-muscle fibers tissues, i.e., blood vessels, fat and connective tissues.

**Table 2.** Histologic characteristics of the quadriceps muscle and their associations with age at death and sex.

|  |  |  | **Model 1** |  | **Model 2** |
|---|---|---|---|---|---|
| **Quadriceps muscle measures** | **Not present, n (%)** | **Mean (SD)** | **Age at death** | **Sex<br>(men vs women)** | **Physical activity** |
|  |  |  | **Estimate (SE), p-value** | **Estimate (SE), p-value** | **Estimate (SE), p-value** |
| **Muscle fiber cross sectional area (µm<sup>2</sup>)** |  |  |  |  |  |
| Type I | 0 | 3352.6 (12764.8) | -0.338 (0.137), 0.014 | -0.626 (1.828), 0.73 | -1.287 (0.874), 0.14 |
| Type II | 2 (0.85) | 1072.2 (845.8) | -0.005 (0.009), 0.55 | 0.736 (0.113), <0.001 | 0.023 (0.052), 0.66 |
| Type IIA | 2 (0.85) | 1295.9 (3164.9) | -0.092 (0.034), 0.007 | 0.393 (0.452), 0.39 | -0.309 (0.222), 0.16 |
| Type IIX | 121 (51.3) | 807.6 (820.1) | -0.004 (0.011), 0.71 | 0.745 (0.147), <0.001 | 0.112 (0.074), 0.13 |
| Mixed Type IIA-IIX | 22 (9.3) | 926.7 (792.6) | -0.007 (0.008), 0.40 | 0.744 (0.107), <0.001 | -0.004 (0.051), 0.93 |
| Mixed Type I-II | 81 (34.3) | 1040.6 (821.5) | 0.001 (0.011), 0.90 | 0.508 (0.145), <0.001 | 0.053 (0.077), 0.50 |
| **Muscle fiber counts (%)** |  |  |  |  |  |
| Type I | 0 | 44.7 (19.5) | 0.205 (0.211), 0.33 | -0.916 (2.827), 0.75 | -0.292 (1.306), 0.82 |
| Type II | 2 (0.85) | 54.2 (19.4) | -0.195 (0.210), 0.36 | 1.182 (2.811), 0.67 | 0.394 (1.299), 0.76 |
| Type IIA | 2 (0.85) | 31.7 (15.1) | -0.003 (0.163), 0.99 | -1.626 (2.183), 0.46 | -0.964 (1.012), 0.34 |
| Type IIX | 121 (51.3) | 2.55 (5.83) | -0.015 (0.062), 0.81 | 2.477 (0.830), 0.003 | 0.310 (0.396), 0.44 |
| Mixed Type IIA-IIX | 22 (9.3) | 20.0 (16.0) | -0.177 (0.173), 0.31 | 0.331 (2.312), 0.89 | 1.049 (1.060), 0.32 |
| Mixed Type I-II | 81 (34.3) | 1.11 (2.14) | -0.010 (0.023), 0.67 | -0.265 (0.310), 0.39 | -0.103 (0.147), 0.48 |
| **Fiber type grouping (%)** |  |  |  |  |  |
| Type I | 117 (49.6) | 11.7 (19.9) | 0.347 (0.215), 0.11 | -1.490 (2.869), 0.60 | 0.834 (1.312), 0.53 |
| Type II | 76 (32.2) | 10.2 (13.5) | -0.021 (0.147), 0.88 | 1.242 (1.963), 0.53 | 1.506 (0.904), 0.097 |
| **Muscle composition components (%)** |  |  |  |  |  |
| Type I | 0 | 40.6 (13.0) | 0.001 (0.001), 0.34 | -0.026 (0.019), 0.17 | -0.009 (0.009), 0.28 |
| Type II | 2 (0.85) | 21.9 (12.9) | -0.003 (0.001), 0.057 | 0.051 (0.018), 0.006 | 0.016 (0.008), 0.062 |
| Type IIA | 2 (0.85) | 13.2 (8.5) | -0.002 (0.001), 0.080 | 0.012 (0.012), 0.33 | 0.006 (0.006), 0.32 |
| Type IIX | 121 (51.3) | 1.0 (2.9) | -0.0001 (0.0003), 0.72 | 0.016 (0.004), <0.001 | 0.002 (0.002), 0.25 |
| Mixed Type IIA-IIX | 22 (9.3) | 7.7 (7.7) | -0.0009 (0.0008), 0.28 | 0.023 (0.011), 0.034 | 0.008 (0.005), 0.12 |
| Mixed Type I-II | 81 (34.3) | 0.5 (1.3) | -0.0000 (0.0001), 0.88 | -0.0009 (0.0019), 0.64 | -0.0002 (0.0009), 0.81 |
| Non-muscle fibers | 0 | 36.9 (8.4) | 0.0013 (0.0009), 0.16 | -0.024 (0.012), 0.048 | -0.006 (0.006), 0.28 |

In separate linear regressions, we examined the associations of predictor terms for **Model 1** (age at death and sex) and **Model 2** (age at death, sex, and physical activity) with the outcomes listed in the left column. **Not present** (second column) indicates the number (percent) of the participants whose quadriceps muscle did not have the characteristics. For example, all participants had type I muscle fibers, but in 121 (51.3%) participants type IIX muscle fibers were not observed.

On average, 36.9% (SD=8.4) of the section area was composed of non-muscle fibers, 40.6% (SD=13.0) was composed of type I muscle fibers, 21.9% (SD=12.9) was composed of type II muscle fibers, including type IIA, IIX, or mixed IIA-IIX fibers, and the remainder, approximately 0.5%, was composed of mixed type I-II muscle fibers. The muscle fibers varied in size, with type I fibers being larger on average than type II fibers. Muscle fiber type grouping was present in more than half of the participants: 50.4% (n=119) had muscle fiber type I grouping and 67.8% (n=160) had type II grouping.

### Age and sex

In separate linear regressions, we examined associations of age at death and sex with the quadriceps muscle histologic characteristics (**Table 2**). Males’ quadriceps muscle sections had relatively larger areas of type II muscle fibers compared with females’, which could be because of more counts of type II fibers, larger size of type II muscle fibers, or both. Further analyses showed that the difference was driven primarily by larger mean type II fiber size in males [mean=1598.9 (SD=1106.7) vs. 860.8 (SD=600.1), p<0.001)], whereas the relative counts of type II fibers did not differ by sex groups (p=0.67). Among type II muscle fibers subtypes, the sex difference in fiber size was observed for type IIX and IIA-IIX but not for type IIA fibers (**Table 2**).

Older age at death was associated with smaller cross-sectional area (CSA) of type I and type IIA muscle fibers, had a borderline association with the percentage of the quadriceps muscle area composed of type II muscle fibers, but not associated with fiber counts (**Table 2**). Neither age nor sex was associated with fiber type I or II grouping.

#### Compositional analysis

For parsimony, we partitioned the quadriceps muscle sections into four areas composed of **a**) type I muscle fibers, **b**) type II muscle fibers, **c**) mixed type I-II muscle fibers, and **d**) area of non-muscle fibers elements. These four components, which were calculated by the ratio of the area of each component to sum of the four areas, comprise the elements of a compositional dataset, since increasing one area is associated with decreasing other(s) (**Figure 2A**) and their summation equals one. Compositional analysis examines whether covariates, such as age at death and sex, are associated with the relative distribution of these components.

**Figure 2.**
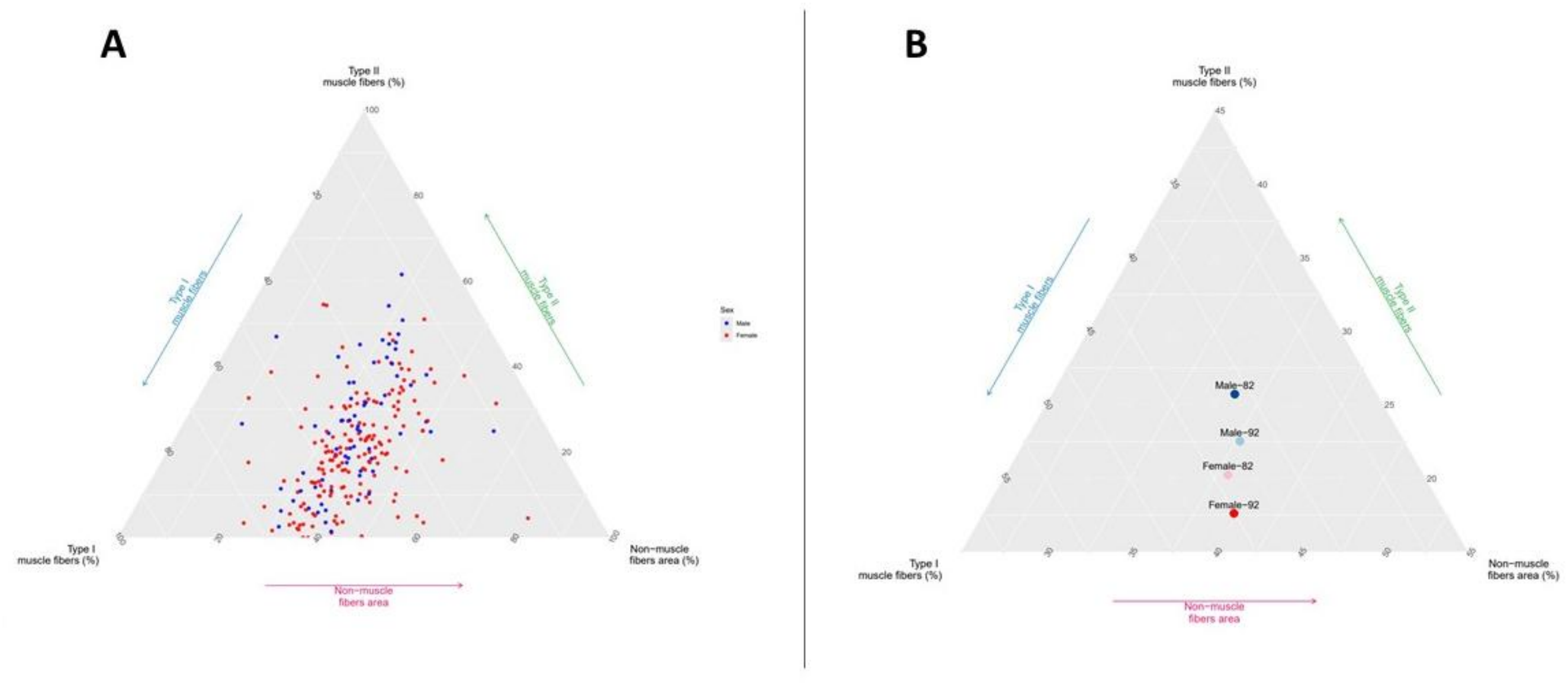
Association of sex and age at death with the four areas of the quadriceps muscle. The ternary plot A shows the distribution of the participants in the simplex space of the four areas of the quadriceps muscle. The ternary plot B shows the composition of the quadriceps muscle of two average males and two average females at age 82 and 92 years old. Younger men had the largest area composed of type II muscle fibers.

In a compositional analysis including age at death and sex, males showed a higher relative proportion of type II fiber area in the quadriceps muscle while older age at death showed a borderline association (p=0.051) with a lower relative proportion of type II fiber area (**Table e-1**). Using the estimated parameters from the compositional analysis, we illustrate the expected muscle composition in four average participants, male and female died at ages 82 and 92 years. The younger male had the highest relative proportion of areas composed of type II muscle fibers followed by the older male, the younger female, and then older female (**Figure 2B**).

### Physical activity

Age and sex are non-modifiable risk factors, so we examined whether the histological characteristics of the quadriceps muscle were related to a common modifiable risk factor, daily physical activity. Daily physical activity was extracted from wrist-worn sensor recordings in 221 participants. The median time interval between the last measurement of daily physical activity and death was 1.60 years (IQR: 0.77 – 3.45), and the mean daily activity counts was 1.22×10^5^ (SD=1.16×10^5^), which is approximately equivalent to 51.8 minutes of walking at 2.5 miles per hour^34^.

In separate linear regressions that controlled for age at death, sex, and the time interval between the last measurement of physical activity and death, higher physical activity showed a borderline association (p=0.062) with a greater relative area composed of type II muscle fibers (**Table 2**). Higher physical activity had a borderline association (p=0.097) with more type II muscle fiber grouping.

#### Compositional analysis

In a compositional analysis with the four muscle components as the outcome and physical activity, age at death, sex, and the time interval between physical activity assessment and death as predictors, more physical activity was associated with larger relative area in the muscle composed of type II muscle fibers (**Table e-1**). Using the estimates derived from the compositional analysis, we compared the composition of quadriceps muscle in two average older adults with high (90^th^ percentile, 2.66×10^5^ activity counts, equivalent to 113 minutes of walking at 2.5 mph) and low (10^th^ percentile, 0.18×10^5^ activity counts, equivalent to 7.6 minutes of walking at 2.5 mph) levels of daily physical activity. As illustrated in **Figure 3**, the relative proportion of the quadriceps muscle composed of type II muscle fibers was 4.8% larger in the individual with the higher level of physical activity.

**Figure 3.**
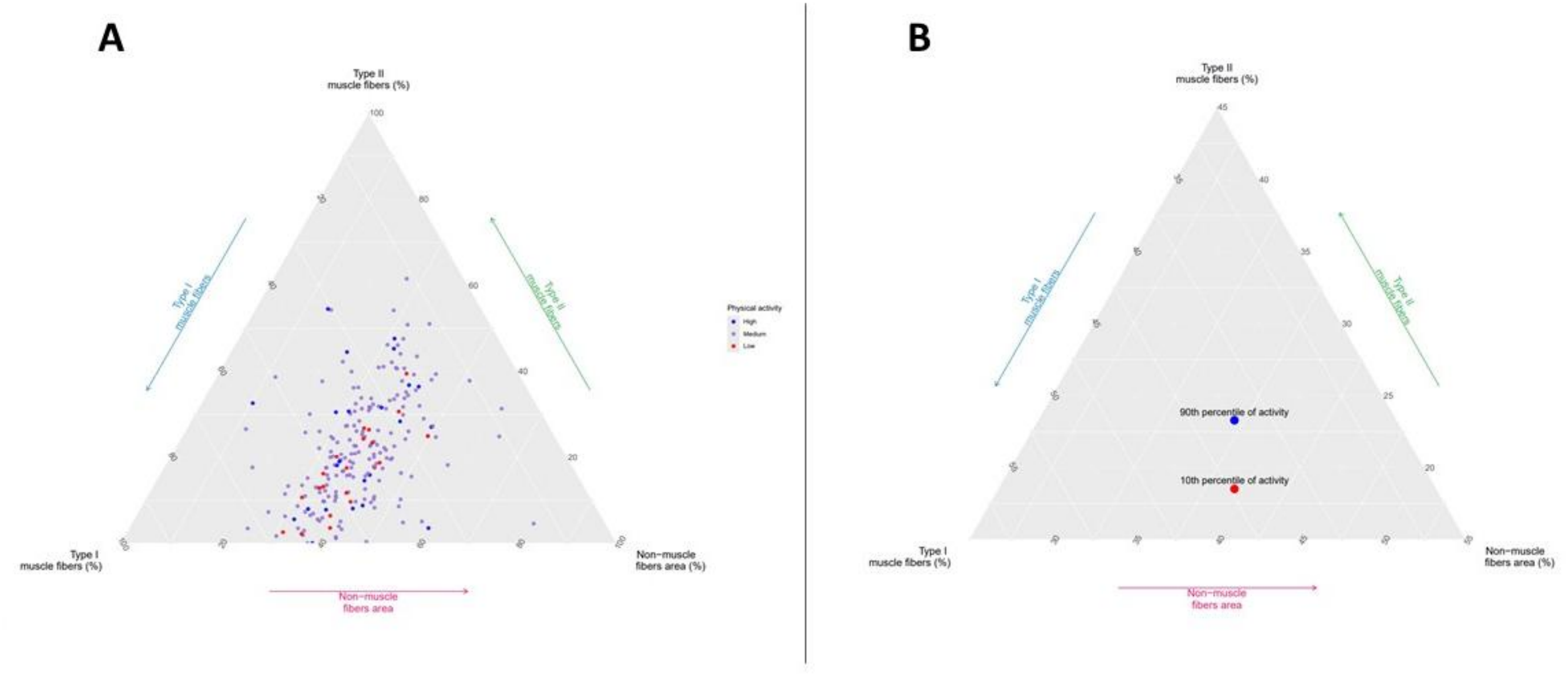
Association of physical activity with the four areas of the quadriceps muscle. The ternary plot A shows the distribution of the participants in the simplex space of the four areas of the quadriceps muscle differentially colored by their level of physical activity. The ternary plot B shows the composition of the quadriceps muscle of two average 92-year-old women with high and low levels of physical activity. Younger woman had a larger area composed of type II muscle fibers.

The association of physical activity with quadriceps muscle composition was materially unchanged after adjusting for vascular risk factors, vascular diseases, low back pain, lower-extremity joint pain, and dementia (**Table e-2**).

### Gait

The median time interval between the last gait function assessment and death was 1.57 years (IQR: 0.80 – 3.01), and the mean gait function at the last visit was 0.71 units (SD=0.22). In separate linear regression models that controlled for age at death, sex, and the time interval between gait function assessment and death, we examined associations of each histological characteristic of the quadriceps muscle with gait function (**Table 3**). A larger area composed of type II muscle fibers and a smaller area composed of type I muscle fibers were associated with better gait function. These associations were appear to be driven by relative fiber counts, rather than by average fiber size. More type II muscle fiber grouping was also associated with better gait.

**Table 3.**
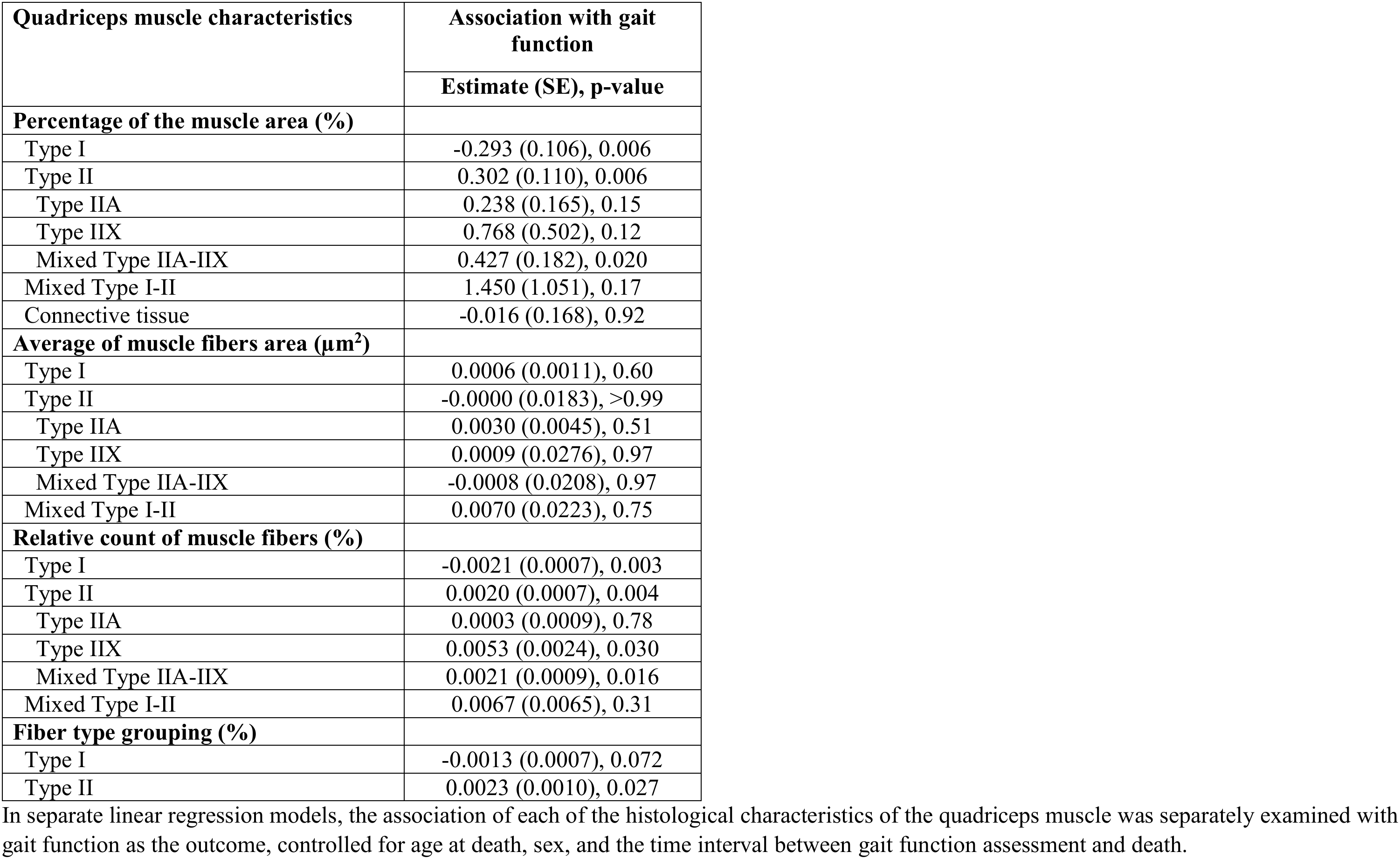
Association of histological characteristics of quadriceps muscle with gait function.

#### Compositional analysis

We used a compositional isotemporal substitution model to examine the four muscle components jointly in relation to gait function. The analysis showed that reallocating area from type II fibers to type I fibers was associated with poorer gait function (**Figure 4**). Compared with the average composition of the 4 muscle areas [type I (40%), type II (22%), mixed (1%), and non-muscle fibers (37%)], reallocating 15% from type II area to type I area was associated with a 0.056-unit worse gait function (95% CI: -0.094 – -0.019), equivalent to 25.5% (95%CI: 8.6%–42.7%) SD of gait function. The reallocation between non-muscle fiber area and type II muscle fiber area was significantly associated with gait function only when reallocating a large portion from type II area to non-muscle fibers area. Specifically, reallocating 15% from type II muscle fibers area to non-muscle fibers area was associated with a 0.052-unit worse gait function (95% CI: -0.103 – -0.002), equivalent to 23.6% (95%CI: 0.91%–46.8%) SD of gait function. The reallocation between non-muscle fiber area with type I muscle fibers area was not associated with gait function

**Figure 4.**
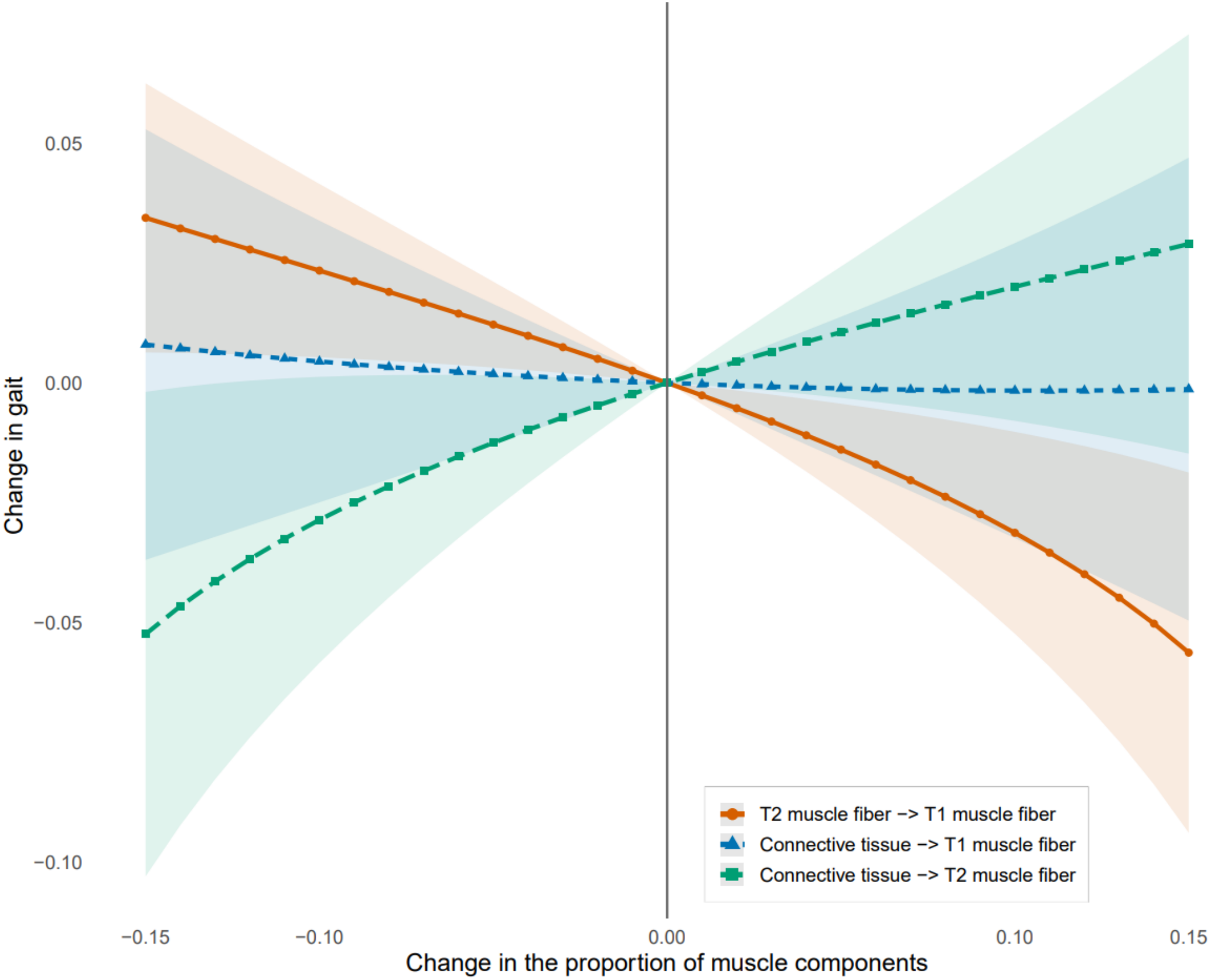
A compositional isotemporal substitution model examining the four areas of the quadriceps muscle in the association with gait function. The only consistently significant change is the one between type I and type II muscle fibers because the confidence interval does not cover zero. The reallocation between connective tissue and type II muscle fibers is significant close to -0.15, when approximately 0.15 (15%) is decreased from the area of type II muscle fibers and added to the area of connective tissues.

In additional analyses, the associations of the quadriceps muscle composition with gait function were materially unchanged after adjusting for vascular risk factors and diseases, low back and lower-extremity joint pain, or dementia (**Table e-3**).

### Quadriceps muscle, ADRD brain pathologies, and gait function

Our previous work showed that gait function declines in older adults after adjusting for ADRD brain pathologies and demographic characteristics including age^17^. Therefore, we tested whether histological characteristics of the quadriceps muscle were associated with gait after adjusting for ADRD pathologies and demographics. Using a compositional isotemporal substitution model, we found that the associations of the quadriceps muscle composition with gait were materially unchanged when adjusted for ADRD brain pathologies (**Table e-3**). Quadriceps muscle composition explained an additional 3.2% of the variance of gait function beyond the 5.8% of variance of gait function explained by ADRD brain pathologies (**Figure 5**, **Table e-4**).

**Figure 5.**
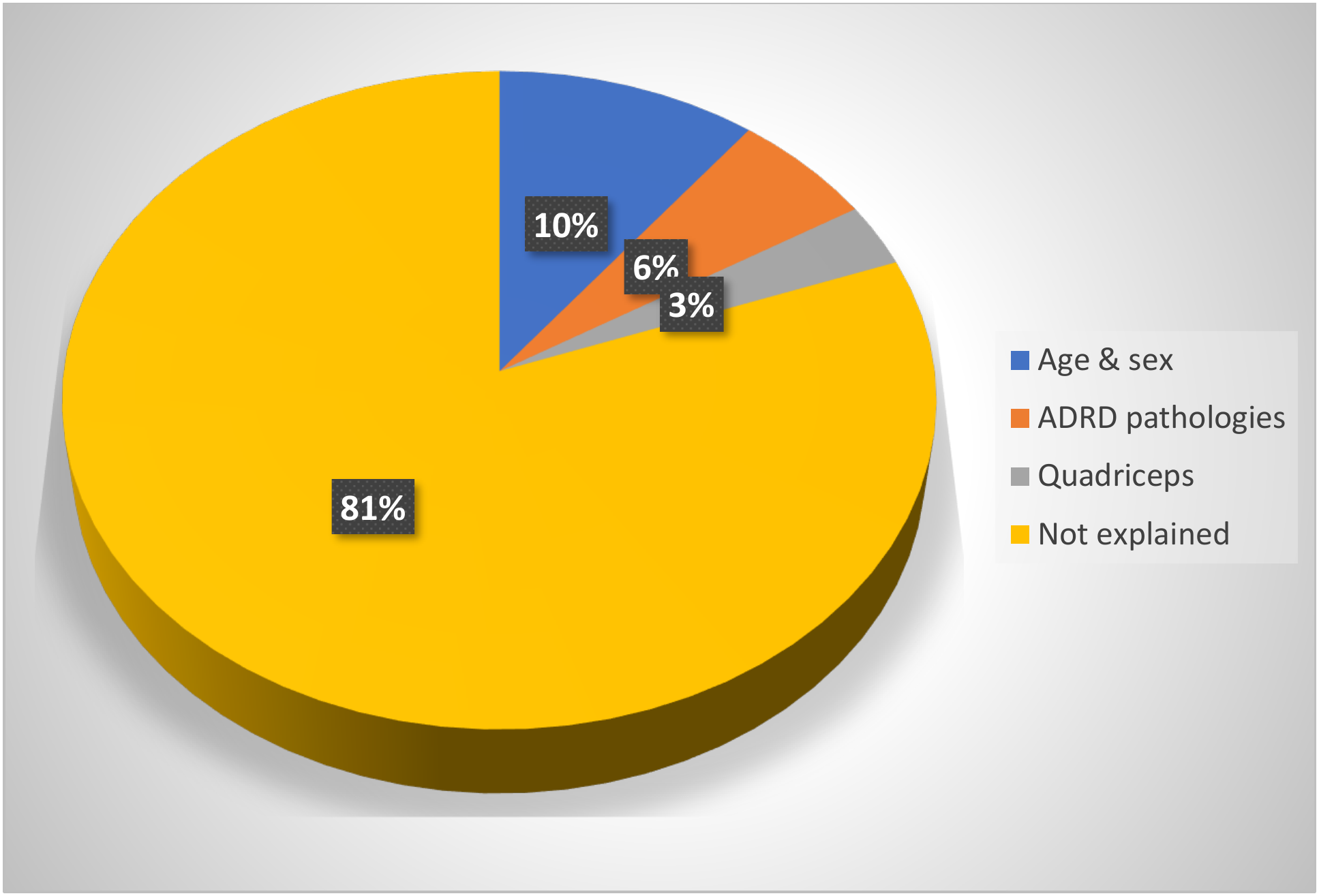
Variance of gait function explained by demographics, Alzheimer’s disease and related dementias (ADRD) pathologies, and the four areas of the quadriceps muscle.

## DISCUSSION

In this clinical-pathological study of approximately 250 older adults, postmortem quadriceps muscle histological characteristics showed that, on average, type I muscle fibers predominated in the quadriceps muscle of older adults. Males had larger areas of the quadriceps muscle composed of type II muscle fibers compared with females. This difference was due mostly to the larger size of males’ type II muscle fibers rather than higher counts. Advancing age was associated with smaller muscle fibers’ cross-sectional areas, particularly for type I fibers and type IIA fibers, but was not associated with muscle fibers counts. Despite the predominance of type I muscle fibers in the quadriceps, more physical activity was associated with larger areas of the quadriceps muscle composed of type II muscle fibers that were associated with better gait in older adults. Differences in the quadriceps muscle composition alone explained more than 3% of the variance of gait function. In contrast, indices of nine ADRD brain pathologies together explained almost 6% of the variance of gait function. While these findings need to be replicated in younger older adults, they support the importance of muscle composition to late-life gait function and underscore the need for further molecular studies to determine the potential efficacy of targeting type II muscle fibers for augmentation of exercise-induced improvement in gait function.

This study extends prior human studies of muscle in several ways. We studied more than 200 participants, whereas most prior studies included fewer than 100 and often fewer than 50. Females comprised more than two thirds of the analytic sample, whereas males predominated in earlier studies. We studied the associations of age, sex, quantitative daily physical activity, and gait function with muscle fibers histological characteristics in individuals from whom indices of ADRD pathologies had been collected. Finally, we used compositional methods to evaluate the 4 tissue components jointly rather than one at a time.

We found decrease in the size of type I and IIA muscle fibers with advancing age, also reported in the prior studies^41–45^. However, we did not find an association between age and muscle fibers counts, which has been reported by some^43^ but not all prior studies^41,42^. While prior studies found associations between aging and muscle histological characteristics by comparing young and older adults, we extended these studies by showing that the age-related decrease in muscle fiber size is observed in very old adults. Multiple pathways may drive age-related muscle fiber atrophy including reduced vascular perfusion, nerve or neuromuscular junction dysfunction, structural changes in the extracellular matrix, mitochondrial dysfunction, and inflammation^46,47^.

Compared with age, sex showed a much stronger association with the histological characteristics of the quadriceps. Males had larger areas of their muscle composed of type II muscle fibers and larger size type II muscle fibers, especially type IIX muscle fibers. This pattern is consistent with a recent meta-analysis with more than 6000 participants showing that males had larger type I and II muscle fibers with a stronger effect size for type II muscle fibers^48^. Prior studies suggest that sex hormones may contribute to sex differences in muscle power^49^ and muscle histology as testosterone administration increased cross sectional area of type II muscle fibers^50^. Moreover, testosterone causes more type II muscle fibers-specific satellite cells^50^, which are internalized by muscle fibers during training or administration of testosterone^51^ and may contribute to increased protein synthesis and larger muscle fiber size. Because the internalized nuclei remain in the muscle^52^ and can resume protein synthesis, they may contribute to enduring sex difference in type II muscle fiber size in older adults^53^.

Few prior studies investigated the association between muscle histologic characteristics and muscle function^54,55^ with limited sample sizes and inadequate separation of type I and II muscle fibers. We found that participants whose quadriceps muscles were composed of more type II muscle fibers had better gait function including faster gait speed. The relative areas of non-muscle fibers compared with muscle fibers did not have a consistent association with gait function, possibly because we did not differentiate different composers of non-muscle fibers tissues including blood vessels, fat and fibrosis. Better gait in older adults had a stronger association with the relative counts of type II versus type I muscle fibers, but muscle fiber size was not related to gait. These novel findings may explain why larger type II fiber size in men does not necessarily translate into lower rates of mobility disability ^56^.

Higher levels of daily physical activity may improve gait because prior studies showed that more physical activity not only causes larger muscle fiber size, specifically type II muscle fibers^57^, but also increase type II muscle fibers counts^58^ through pathways such as stimulation of satellite cells specific for type II muscle fibers^59^ or angiogenesis of capillaries around the muscle fibers^58^. Because more muscle fiber type II grouping was also associated with better gait in our study, another possible explanation is that factors such as long-term regular exercise may lead to remodeling^60^ of the muscle fibers from type I to type II muscle fibers through reinnervation^61^ via fast motor neurons^62^. Fiber type grouping is a classic histologic correlate of reinnervation; when surviving fast motor neurons reinnervate denervated fibers, territories can skew toward type II profiles, aligning with our finding that greater type II proportion and grouping are associated with better gait performance^63–65^. This model predicts that direct markers of denervation (axon loss, NMJ fragmentation) and reinnervation (sprouting indices) in quadriceps or peripheral nerves should co vary with type II grouping and gait in late life, a prediction for investigation in future studies.

In prior work we showed that motor decline in old age is not accounted for by increasing age alone ^17^. Prior studies also showed that although ADRD pathologies in the brain are associated with declining motor function in older adults^5,16–19^, they explain less than 10% of the variance of motor decline^19^. Therefore, pathologies and molecular changes in other motor-related tissues in the CNS^29^, peripheral nervous system^66^, and musculoskeletal structures that comprise the distributed motor pathways must be examined to more fully account for declining motor function in aging adults. The current findings extend prior brain autopsy results in this cohort by finding that quantifying skeletal muscle histologic features explains additional variance of gait beyond the variance accounted for by brain ADRD pathologies. These findings reinforce the concept that both upper and lower motor neuron structures (motor neuron, axon, NMJ, and myofiber ensemble) are critical determinants of mobility in aging adults. Since no treatments for ADRD brain pathologies are currently available, these results emphasize the need for studies focusing on targeted treatments for muscle degeneration to improve gait and prevent mobility disability in elders.

Several limitations must be considered when interpreting these findings. This was a cross-sectional study, which precludes causal inference. Because the age range was relatively narrow, participants were mostly in their 9^th^ and 10th decades of life, we might have limited power to detect some of the aging effects previously reported, such as type I muscle fiber grouping, which were derived by comparing adults 20-40 years old with those older than 60 years^41^. The analytic sample was almost completely composed of non-Latino White participants with relatively high educational attainment than the average American elders, which requires findings be tested in more diverse samples for generalization. We did not have detailed information about the severity or the types of physical exercise to test if the association of physical activity with muscle histology varied by those parameters. We examined only the quadriceps muscle, and the findings may not directly generalize to other muscles because of the varied histological properties across muscles^67,68^. We did not have information about mobility status during the last months before death, which might have affected our findings considering the association of disuse and inactivity with muscle fiber type II atrophy^69^. Because Nonetheless, this study has several notable strengths including the relatively large sample size, examining physical activity and gait function of the same individuals in relation to the histologic characteristics of their muscles, the ability to compare muscle findings with ADRD brain pathologies in relation to gait, and the use of compositional methods to analyze muscle components distribution in relation to physical activity and gait.

## Data Availability

To access the data qualified investigators should submit a short proposal including a brief description of the study and a research plan and should sign a data use agreement.

http://www.radc.rush.edu

## DECLARATIONS

### Ethics approval and consent to participate

Written informed consents were obtained from all the participants, including signed Anatomical Gift Act for brain donation. The MAP study was approved by a Rush University Medical Center Institutional Review Board with the IRB approval number of L86121802.

### Consent for publication

Not applicable.

## Availability of data and materials

All data generated and used for the current study are available via the Rush Alzheimer’s Disease Center Research Resource Sharing Hub, www.radc.rush.edu. To obtain data, qualified investigators should fill a short application describing the premises of their research and an overview of the analytic plans.

## Authors’ contributions

S.O. conceptualized the study, designed the analyses, interpreted the findings, and wrote the first draft. S.L. collected quantified histological characteristics of the quadriceps muscle and participated in preparing Figure 1 and Figure e-1. J.Y. supervised and did the statistical analyses, prepared or supervised making Figures 2-5, and critically reviewed the manuscript. J.A.S. supervised collecting Alzheimer’s disease and related dementias pathologies in the brain, and critically reviewed the manuscript. D.A.B. is the principal investigator of the parent cohort study, the Rush Memory and Aging Project, critically reviewed the manuscript, and contributed in providing fund for conducting the study. O.D. supervised dissection of the quadriceps muscle samples and collecting quantified histological characteristics of the quadriceps muscle, prepared Figure 1 and Figure e-1, and critically reviewed the manuscript. A.S.B. participated in seeking funds to conduct the study, in conceptualizing the study, in interpretation of the findings, and in critically reviewing the manuscript. All authors reviewed the manuscript.

## Acknowledgement

We acknowledge MAP participants who generously donated their time and were willing to allow autopsies to advance our knowledge about aging-related phenotypes and their associated risk factors and mechanisms. We also acknowledge the meticulous work of our colleagues at Rush Alzheimer’s Disease Center who participated in collecting, handling, and analyzing the study data.

## REFERENCES

1. Courtney-Long EA, Carroll DD, Zhang QC, et al. Prevalence of Disability and Disability Type Among Adults--United States, 2013. MMWR Morb Mortal Wkly Rep. 2015;64:777–783.

2. Sullivan KJ, Ranadive R, Su D, et al. Imaging-based indices of Neuropathology and gait speed decline in older adults: the atherosclerosis risk in communities study. Brain Imaging Behav. 2021;15:2387–2396.

3. Tian Q, Resnick SM, Bilgel M, Wong DF, Ferrucci L, Studenski SA. β-Amyloid burden predicts lower extremity performance decline in cognitively unimpaired older adults. J Gerontol A Biol Sci Med Sci. 2017;72:716–723.

4. Wennberg AMV, Lesnick TG, Schwarz CG, et al. Longitudinal Association Between Brain Amyloid-Beta and Gait in the Mayo Clinic Study of Aging. J Gerontol A Biol Sci Med Sci. 2018;73:1244–1250.

5. Rosso AL, Studenski SA, Longstreth WT, Brach JS, Boudreau RM, Rosano C. Contributors to Poor Mobility in Older Adults: Integrating White Matter Hyperintensities and Conditions Affecting Other Systems. J Gerontol A Biol Sci Med Sci. 2017;72:1246–1251.

6. Cruz-Jentoft AJ, Sayer AA. Sarcopenia. The Lancet. 2019;393:2636–2646.

7. Cruz-Jentoft AJ, Bahat G, Bauer J, et al. Sarcopenia: revised European consensus on definition and diagnosis. Age Ageing. 2019;48:16–31.

8. Wang DXM, Yao J, Zirek Y, Reijnierse EM, Maier AB. Muscle mass, strength, and physical performance predicting activities of daily living: a meta-analysis. J Cachexia Sarcopenia Muscle. 2020;11:3–25.

9. Takakusaki K. Functional neuroanatomy for posture and gait control. J Mov Disord. 2017;10:1–17.

10. Beaudart C, McCloskey E, Bruyère O, et al. Sarcopenia in daily practice: assessment and management. BMC Geriatr. 2016;16:170.

11. Naruse M, Trappe S, Trappe TA. Human skeletal muscle-specific atrophy with aging: a comprehensive review. J Appl Physiol Bethesda Md 1985. 2023;134:900–914.

12. Sales WB, Mâcedo SGGF, Gonçalves RSDSA, et al. Use of electrical bioimpedance in the assessment of sarcopenia in the older aldults: A scoping review. J Bodyw Mov Ther. 2024;39:373–381.

13. Zhao R, Li X, Jiang Y, et al. Evaluation of Appendicular Muscle Mass in Sarcopenia in Older Adults Using Ultrasonography: A Systematic Review and Meta-Analysis. Gerontology. 2022;68:1174–1198.

14. Lexell J. Human aging, muscle mass, and fiber type composition. J Gerontol A Biol Sci Med Sci. 1995;50 Spec No:11–16.

15. Tanganelli F, Meinke P, Hofmeister F, et al. Type-2 muscle fiber atrophy is associated with sarcopenia in elderly men with hip fracture. Exp Gerontol. 2021;144:111171.

16. Buchman AS, Wang T, Yu L, Leurgans SE, Schneider JA, Bennett DA. Brain pathologies are associated with both the rate and variability of declining motor function in older adults. Acta Neuropathol (Berl). 2020;140:587–589.

17. Oveisgharan S, Wang T, Barnes LL, Schneider JA, Bennett DA, Buchman AS. The time course of motor and cognitive decline in older adults and their associations with brain pathologies: a multicohort study. Lancet Healthy Longev. 2024;5:e336–e345.

18. Oveisgharan S, Yu L, Agrawal S, et al. Relation of Motor Impairments to Neuropathologic Changes of Limbic-Predominant Age-Related TDP-43 Encephalopathy in Older Adults. Neurology. 2023;101:e1542–e1553.

19. Oveisgharan S, Yu L, Wang T, Schneider JA, Bennett DA, Buchman AS. Neurodegenerative and cerebrovascular brain pathologies are differentially associated with declining grip strength and gait In older adults. J Gerontol A Biol Sci Med Sci. 2023;78:504–513.

20. Bonilla HJ, Messi ML, Sadieva KA, Hamilton CA, Buchman AS, Delbono O. Semiautomatic morphometric analysis of skeletal muscle obtained by needle biopsy in older adults. GeroScience. 2020;42:1431–1443.

21. Legland D, Arganda-Carreras I, Andrey P. MorphoLibJ: integrated library and plugins for mathematical morphology with ImageJ. Bioinformatics. Oxford, England; 2016;32:3532–3534.

22. Kapasi A, Poirier J, Hedayat A, et al. High-throughput digital quantification of Alzheimer disease pathology and associated infrastructure in large autopsy studies. J Neuropathol Exp Neurol. 2023;82:976–986.

23. Hyman BT, Phelps CH, Beach TG, et al. National Institute on Aging-Alzheimer’s Association guidelines for the neuropathologic assessment of Alzheimer’s disease. Alzheimers Dement J Alzheimers Assoc. 2012;8:1–13.

24. Agrawal S, Yu L, Kapasi A, et al. Limbic-predominant age-related TDP-43 encephalopathy neuropathologic change and microvascular pathologies in community-dwelling older persons. Brain Pathol. 2021;31:e12939.

25. Oveisgharan S, Grodstein F, Evia AM, et al. Association of age-related neuropathologic findings at autopsy with a claims-based epilepsy diagnosis in older adults. Neurology. 2024;102:e209172.

26. Agrawal S, Leurgans SE, Nag S, et al. Effects of Cerebrovascular and Lewy Body Pathology on Parkinsonian Signs in Community-Dwelling Older Adults. Neurology. 2023;101:e754–e763.

27. Buchman AS, Yu L, Oveisgharan S, et al. Associations of pathologic Parkinson’s disease (PD) and co-pathologies with cognitive decline and progression of parkinsonian signs in decedents with subclinical disease. Acta Neuropathol (Berl). 2025;151:1.

28. Oveisgharan S, Dawe RJ, Yu L, et al. Frequency and underlying pathology of pure vascular cognitive impairment. JAMA Neurol. 2022;79:1277–1286.

29. Oveisgharan S, Agrawal S, Yu L, et al. Arteriolosclerosis in CNS Tissues Outside the Cerebrum and Late-Life Motor Impairment. Stroke. Epub 2025 Apr 29.

30. Oveisgharan S, Yu L, Barnes LL, et al. Association of statins with cerebral atherosclerosis and incident parkinsonism in older adults. Neurology. Epub 2022 Mar 23.:10.1212/WNL.0000000000200182.

31. Oveisgharan S, Kim N, Agrawal S, et al. Brain and spinal cord arteriolosclerosis and its associations with cerebrovascular disease risk factors in community-dwelling older adults. Acta Neuropathol (Berl). 2023;145:219–233.

32. Oveisgharan S, Yu L, Yang J, et al. Cortical gray matter proteins associated with cerebral amyloid angiopathy in community-dwelling older adults: An autopsy study. Neurology. 2025;105:e214024.

33. Oveisgharan S, Wang T, Hausdorff JM, Bennett DA, Buchman AS. Motor and Nonmotor Measures and Declining Daily Physical Activity in Older Adults. JAMA Netw Open. 2024;7:e2432033.

34. Heil DP. Predicting activity energy expenditure using the Actical activity monitor. Res Q Exerc Sport. 2006;77:64–80.

35. Boyle PA, Wilson RS, Buchman AS, et al. Lower extremity motor function and disability in mild cognitive impairment. Exp Aging Res. 2007;33:355–371.

36. McKhann GM, Knopman DS, Chertkow H, et al. The diagnosis of dementia due to Alzheimer’s disease: recommendations from the National Institute on Aging-Alzheimer’s Association workgroups on diagnostic guidelines for Alzheimer’s disease. Alzheimers Dement. 2011;7:263–269.

37. Dumuid D, Pedišić Ž, Palarea-Albaladejo J, Martín-Fernández JA, Hron K, Olds T. Compositional Data Analysis in Time-Use Epidemiology: What, Why, How. Int J Environ Res Public Health. 2020;17:2220.

38. Gloor GB, Reid G. Compositional analysis: a valid approach to analyze microbiome high-throughput sequencing data. Can J Microbiol. 2016;62:692–703.

39. Smithson M, Broomell SB. Compositional data analysis tutorial. Psychol Methods. Epub 2022 Jan 31.

40. Dumuid D, Pedišić Ž, Stanford TE, et al. The compositional isotemporal substitution model: A method for estimating changes in a health outcome for reallocation of time between sleep, physical activity and sedentary behaviour. Stat Methods Med Res. 2019;28:846–857.

41. Lexell J. Human aging, muscle mass, and fiber type composition. J Gerontol A Biol Sci Med Sci. 1995;50 Spec No:11–16.

42. Bougea A, Papadimas G, Papadopoulos C, et al. An Age-Related Morphometric Profile of Skeletal Muscle in Healthy Untrained Women. J Clin Med. 2016;5:97.

43. Nilwik R, Snijders T, Leenders M, et al. The decline in skeletal muscle mass with aging is mainly attributed to a reduction in type II muscle fiber size. Exp Gerontol. 2013;48:492–498.

44. Poggi P, Marchetti C, Scelsi R. Automatic morphometric analysis of skeletal muscle fibers in the aging man. Anat Rec. 1987;217:30–34.

45. Nikolić M, Malnar-Dragojević D, Bobinac D, Bajek S, Jerković R, Soić-Vranić T. Age-related skeletal muscle atrophy in humans: an immunohistochemical and morphometric study. Coll Antropol. 2001;25:545–553.

46. Granic A, Suetterlin K, Shavlakadze T, Grounds MD, Sayer AA. Hallmarks of ageing in human skeletal muscle and implications for understanding the pathophysiology of sarcopenia in women and men. Clin Sci Lond Engl 1979. 2023;137:1721–1751.

47. Sayer AA, Cooper R, Arai H, et al. Sarcopenia. Nat Rev Dis Primer. 2024;10:68.

48. James JJ, Mellow ML, Bueckers EP, et al. Sex differences in human skeletal muscle fiber types and the influence of age, physical activity, and muscle group: A systematic review and meta-analysis. Physiol Rep. 2025;13:e70616.

49. Hunter SK, Angadi SS, Bhargava A, et al. The Biological Basis of Sex Differences in Athletic Performance: Consensus Statement for the American College of Sports Medicine. Med Sci Sports Exerc. 2023;55:2328–2360.

50. Horwath O, Apró W, Moberg M, et al. Fiber type-specific hypertrophy and increased capillarization in skeletal muscle following testosterone administration in young women. J Appl Physiol. Bethesda, Md. : 1985; 2020;128:1240–1250.

51. Sinha-Hikim I, Artaza J, Woodhouse L, et al. Testosterone-induced increase in muscle size in healthy young men is associated with muscle fiber hypertrophy. Am J Physiol Endocrinol Metab. 2002;283:E154–164.

52. Bruusgaard JC, Johansen IB, Egner IM, Rana ZA, Gundersen K. Myonuclei acquired by overload exercise precede hypertrophy and are not lost on detraining. Proc Natl Acad Sci U S A. 2010;107:15111–15116.

53. Wu FCW, Tajar A, Pye SR, et al. Hypothalamic-pituitary-testicular axis disruptions in older men are differentially linked to age and modifiable risk factors: the European Male Aging Study. J Clin Endocrinol Metab. 2008;93:2737–2745.

54. Mastavičiūtė A, Kilaitė J, Petroška D, Laurinavičius A, Tamulaitienė M, Alekna V. Associations between Physical Function, Bone Density, Muscle Mass and Muscle Morphology in Older Men with Sarcopenia: A Pilot Study. Medicina (Mex). Kaunas, Lithuania; 2021;57:156.

55. Nicklas BJ, Leng I, Delbono O, et al. Relationship of physical function to vastus lateralis capillary density and metabolic enzyme activity in elderly men and women. Aging Clin Exp Res. 2008;20:302–309.

56. Lundgren-Lindquist B, Jette AM. Mobility disability among elderly men and women in Sweden. Int Disabil Stud. 1990;12:1–5.

57. Kosek DJ, Kim J-S, Petrella JK, Cross JM, Bamman MM. Efficacy of 3 days/wk resistance training on myofiber hypertrophy and myogenic mechanisms in young vs. older adults. J Appl Physiol. Bethesda, Md. : 1985; 2006;101:531–544.

58. Moro T, Brightwell CR, Volpi E, Rasmussen BB, Fry CS. Resistance exercise training promotes fiber type-specific myonuclear adaptations in older adults. J Appl Physiol. Bethesda, Md. : 1985; 2020;128:795–804.

59. Verdijk LB, Gleeson BG, Jonkers RAM, et al. Skeletal muscle hypertrophy following resistance training is accompanied by a fiber type-specific increase in satellite cell content in elderly men. J Gerontol A Biol Sci Med Sci. 2009;64:332–339.

60. Kelly NA, Hammond KG, Bickel CS, Windham ST, Tuggle SC, Bamman MM. Effects of aging and Parkinson’s disease on motor unit remodeling: influence of resistance exercise training. J Appl Physiol. Bethesda, Md. : 1985; 2018;124:888–898.

61. Coletti C, Acosta GF, Keslacy S, Coletti D. Exercise-mediated reinnervation of skeletal muscle in elderly people: An update. Eur J Transl Myol. 2022;32:10416.

62. Stifani N. Motor neurons and the generation of spinal motor neuron diversity. Front Cell Neurosci. 2014;8:293.

63. Delbono O, Wang Z-M, Messi ML. The rise and deceleration of neuronal excitability in aging and Alzheimer’s disease: Mechanisms, implications, and therapeutic targets. Ageing Res Rev. 2026;114:102999.

64. Delbono O. Neural control of aging skeletal muscle. Aging Cell. 2003;2:21–29.

65. Messi ML, Li T, Wang Z-M, Marsh AP, Nicklas B, Delbono O. Resistance Training Enhances Skeletal Muscle Innervation Without Modifying the Number of Satellite Cells or their Myofiber Association in Obese Older Adults. J Gerontol A Biol Sci Med Sci. 2016;71:1273–1280.

66. Oveisgharan S, Collin A, Yang J, et al. Morphometric characteristics of tibial nerve and their relationship with age. Brain Commun. 2025;7:fcaf267.

67. Edgerton VR, Smith JL, Simpson DR. Muscle fibre type populations of human leg muscles. Histochem J. 1975;7:259–266.

68. Tesch PA, Karlsson J. Muscle fiber types and size in trained and untrained muscles of elite athletes. J Appl Physiol. Bethesda, Md. : 1985; 1985;59:1716–1720.

69. Hvid LG, Ortenblad N, Aagaard P, Kjaer M, Suetta C. Effects of ageing on single muscle fibre contractile function following short-term immobilisation. J Physiol. 2011;589:4745–4757.

