## Supplemental File for "Histological characteristics of the quadriceps muscle and their associations with gait in older adults"

| **Content** | **Pages** |
|---|---|
| **Figure e-1. Cross sections of the quadriceps muscles enriched for different types of muscle fibers.** | **2** |
| **Table e-1. Association of age at death, sex, and physical activity with quadriceps muscle composition.** | **3** |
| **Table e-2. Association of physical activity with quadriceps muscle composition after controlling for covariates.** | **4** |
| **Table e-3. Association of quadriceps muscle composition with gait function after controlling for covariates.** | **5** |
| **Table e-4. Variance of gait function explained by quadriceps muscle composition and Alzheimer's disease and related dementias (ADRD) pathologies.** | **6** |

**Figure e-1. Cross sections of the quadriceps muscles enriched for different types of muscle fibers.**

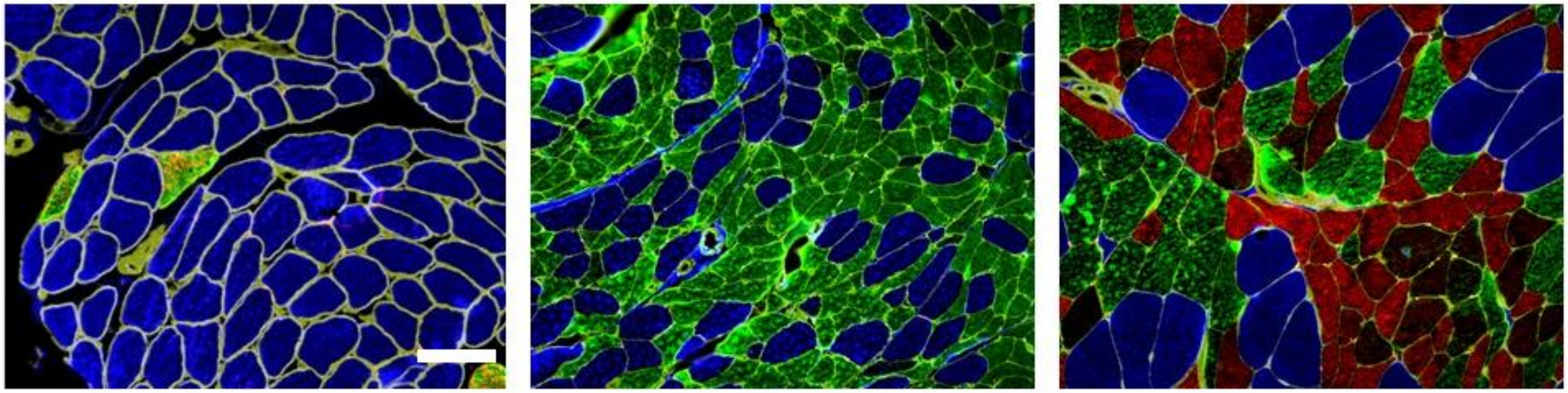

The figure shows muscles enriched in type I, type IIA, and type IIX fibers, from left to right. Scale bar = 100 µm.

**Table e-1. Association of age at death, sex, and physical activity with quadriceps muscle composition.**

| **Model terms** | **Model 1 Age and sex** | **Model 2 Physical activity** |
|---|---|---|
| | **Estimate (SE), p-value** | |
| Intercept | 5.924 (0.166), <0.001 | 5.940 (0.238), <0.001 |
| Age at death | 0.017 (0.023), 0.47 | 0.031 (0.025), 0.22 |
| Sex (male vs. Female) | 0.273 (0.312), 0.38 | 0.448 (0.319), 0.16 |
| Percentage area of type I muscle fibers/percentage area of mixed muscle fibers | 0.066 (0.067), 0.33 | 0.072 (0.096), 0.45 |
| Percentage area of type II muscle fibers/percentage area of mixed muscle fibers | -0.818 (0.067), <0.001 | -0.776 (0.096), <0.001 |
| Percentage area of non-muscle fibers/percentage area of mixed muscle fibers | **REF** | **REF** |
| Age at death×(percentage area of type I muscle fibers/percentage area of mixed muscle fibers) | -0.002 (0.009), 0.84 | 0.001 (0.010), 0.91 |
| Age at death×(percentage area of type II muscle fibers/percentage area of mixed muscle fibers) | -0.018 (0.009), 0.051 | -0.012 (0.010), 0.24 |
| Sex×(percentage area of type I muscle fibers/percentage area of mixed muscle fibers) | -0.013 (0.126), 0.92 | -0.021 (0.128), 0.87 |
| Sex×(percentage area of type II muscle fibers/percentage area of mixed muscle fibers) | 0.303 (0.126), 0.016 | 0.238 (0.128), 0.063 |
| Physical activity | NA | 0.317 (0.143), 0.028 |
| Physical activity×(percentage area of type I muscle fibers/percentage area of mixed muscle fibers) | NA | 0.001 (0.057), 0.98 |
| Physical activity×(percentage area of type II muscle fibers/percentage area of mixed muscle fibers) | NA | 0.118 (0.059), 0.045 |
| Time from physical activity assessment to death | NA | -0.050 (0.065), 0.45 |
| Time_physical activity×(percentage area of type I muscle fibers/percentage area of mixed muscle fibers) | NA | -0.004 (0.026), 0.87 |
| Time_physical activity×(percentage area of type II muscle fibers/percentage area of mixed muscle fibers) | NA | -0.006 (0.026), 0.81 |

The estimates, SE, and p-values are derived from two separate mixed effects models, with the fixed effects listed in the left column and a random intercept effect for each participant. The outcome were the logarithm transformation of the ratios of the percentage areas of type I, type II, and non-muscle fibers to percent area of the mixed muscle fibers.

**Table e-2. Association of physical activity with quadriceps muscle composition after controlling for covariates.**

| Model terms | Vascular risk factors | Body mass index | Vascular diseases | Pain | Dementia |
|---|---|---|---|---|---|
| | Estimate (SE), p-value | | | | |
| Percentage area of type I muscle fibers/percentage area of mixed muscle fibers | 0.095 (0.134), 0.48 | 0.069 (0.095), 0.47 | 0.059 (0.107), 0.58 | 0.036 (0.105), 0.74 | 0.079 (0.100), 0.43 |
| Percentage area of type II muscle fibers/percentage area of mixed muscle fibers | -1.007 (0.135), <0.001 | -0.761 (0.096), <0.001 | -0.806 (0.107), <0.001 | -0.768 (0.106), <0.001 | -0.790 (0.100), <0.001 |
| Percentage area of non-muscle fibers/percentage area of mixed muscle fibers | **REF** | **REF** | **REF** | **REF** | **REF** |
| Physical activity | 0.314 (0.143), 0.030 | 0.309 (0.143), 0.032 | 0.310 (0.143), 0.031 | 0.294 (0.145), 0.044 | 0.298 (0.148), 0.046 |
| Physical activity×(percentage area of type I muscle fibers/percentage area of mixed muscle fibers) | 0.001 (0.057), 0.99 | 0.000 (0.057), >0.99 | 0.002 (0.058), 0.98 | -0.003 (0.058), 0.96 | -0.008 (0.059), 0.90 |
| Physical activity×(percentage area of type II muscle fibers/percentage area of mixed muscle fibers) | 0.123 (0.058), 0.035 | 0.124 (0.058), 0.035 | 0.120 (0.059), 0.042 | 0.119 (0.060), 0.047 | 0.135 (0.061), 0.027 |

The estimates, SE, and p-values are derived from five separate mixed effects models, with the fixed effects terms related to physical activity listed in the left column. Other fixed effects terms were age at death, sex, time from the last visit of physical activity measurement to death, and one of the covariates listed in the five right columns, and the interaction of these terms with the two percentage area of type I and type II muscle fibers divided by percent area of mixed muscle fibers. The outcome were the logarithm transformation of the ratios of the percentage areas of type I, type II, and non-muscle fibers to percent area of the mixed muscle fibers.

**Table e-3. Association of quadriceps muscle composition with gait function after controlling for covariates.**

| The transformed muscle components | Reference | Vascular risk factor | Body mass index | Vascular diseases | Pain | Dementia | ADRD pathologies |
|---|---|---|---|---|---|---|---|
| | Estimate (SE), p-value | | | | | | |
| Variable 1 | 0.014 (0.033), 0.66 | 0.015 (0.033), 0.65 | 0.016 (0.032), 0.61 | 0.016 (0.032), 0.61 | 0.015 (0.032), 0.63 | 0.014 (0.031), 0.64 | 0.003 (0.034), 0.93 |
| Variable 2 | -0.049 (0.021), 0.021 | -0.049 (0.021), 0.021 | -0.053 (0.021), 0.011 | -0.050 (0.020), 0.014 | -0.051 (0.020), 0.012 | -0.051 (0.020), 0.011 | -0.053 (0.021), 0.014 |
| Variable 3 | 0.010 (0.023), 0.67 | 0.010 (0.023), 0.66 | 0.013 (0.023), 0.57 | 0.015 (0.022), 0.50 | 0.012 (0.022), 0.58 | 0.011 (0.022), 0.61 | 0.003 (0.024), 0.89 |

The formulas for transforming the four areas composing the quadriceps muscle and bringing them from a simplex to a real space are as follows:

Variable 1= ln(square root(area of type muscle fibers×area of type II muscle fibers)/square root(area of mixed muscle fibers×non-muscle fibers area)).

Variable 2= $0.5^{1/2}$×ln(area of type I muscle fibers/area of type II muscle fibers).

Variable 3= $0.5^{1/2}$×ln(area of mixed muscle fibers/area of non-muscle fibers).

In seven linear regression models, the associations of the four areas of the quadriceps muscle with gait function was examined. The outcomes of the models were gait function in the last visit. The model terms of the reference model were the three transformed variables, age at death, sex, and the time interval between gait assessment and death. The following covariates were successively added to the reference model's terms: a summary measure of vascular risk factors (indicating the number of hypertension, diabetes mellitus, and smoking present in a participant), body mass index, a summary measure of vascular diseases (indicating the number of stroke, myocardial infarction, heart failure, and lower extremities claudication present in a participant), low back pain and pain the lower extremities' joints, and dementia. The last model included the reference model terms and the brain Alzheimer's disease and related dementias (ADRD) pathologies: amyloid-β load, tau tangle density, Parkinson's disease pathology, transactive response DNA-binding protein 43 aggregates, hippocampal sclerosis, macoinfarcts, microinfarcts, atherosclerosis, arteriolosclerosis, and cerebral amyloid angiopathy. The analyses indicate that the association of the four areas of the quadriceps muscle with gait did not change after controlling for further covariates or controlling for ADRD pathologies.

**Table e-4. Variance of gait function explained by quadriceps muscle composition and Alzheimer's disease and related dementias (ADRD) pathologies.**

| Model | Model terms | $R^2$ |
|---|---|---|
| **1** | **Age at death, sex, time between gait assessment and death** | 0.105 |
| **2** | **Model 1 terms, the transformed variables derived from the four areas of the quadriceps muscle** | 0.136 |
| **3** | **Model 1 terms, ADRD pathologies** | 0.163 |
| **4** | **Model 2 terms, ADRD pathologies** | 0.195 |

In four separate linear regressions, the associations of the model terms, listed in the middle column, were examined with gait function, as the outcome. The transformed variables are described in the text and at the bottom of Table e-3.
